# Associations between state-level healthcare access and COVID-19 case trajectories in the United States

**DOI:** 10.1101/2020.07.04.20146100

**Authors:** Shelley H. Liu, Bian Liu, Agnes Norbury, Yan Li

## Abstract

**Introduction:** We conducted an ecological study to determine if state-level healthcare access is associated with trajectories of daily reported COVID-19 cases in the United States. Our focus is on trajectories of daily reported COVID-19 cases, rather than cumulative cases, as trajectories help us identify trends in how the pandemic naturally develops over time, and study the shapes of the curve in different states.

**Methods:** We analyzed data on daily reported confirmed and probable COVID-19 cases from January 21 to June 16, 2020 in 50 states, adjusted for the population size of each state. Cluster analysis for time-series data was used to split the states into clusters that have distinct trajectories of daily cases. Differences in socio-demographic characteristics and healthcare access between clusters were tested. Adjusted models were used to determine if healthcare access is associated with reporting a high trajectory of COVID-19 cases.

**Results:** Two clusters of states were identified. One cluster had a high trajectory of population-adjusted COVID-19 cases, and comprised of 19 states, including New York and New Jersey. The other cluster of states (n=31) had a low trajectory of population-adjusted COVID-19 cases.

There were significantly more Black residents (p=0.027) and more nursing facility residents (p=0.001) in states reporting high trajectory of COVID-19 cases. States reporting a high trajectory of COVID-19 cases also had fewer uninsured persons (p=0.005), fewer persons who reported having to forgo medical care due to cost (p=0.016), more registered physicians (p=0.002) and more nurses (p=0.03), higher health spending per capita (p=0.01), fewer residents in Health Professional Shortage Areas per 100,000 population (p=0.027), and higher adoption of Medicaid Expansion (p=0.05).

In adjusted models, a higher proportion of uninsured persons (OR: 0.51 [0.25-0.85]; p=0.032), higher proportion of patients who had to forgo medical care due to cost (OR: 0.55 [0.28-0.95]; p=0.048), and no adoption of Medicaid expansion (OR: 0.05 [0 – 0.59]; p=0.04), were associated with reporting a low trajectory of COVID-19 cases.

**Conclusion:** Our findings from adjusted models suggest that healthcare access can partially explain variations in COVID-19 case trajectories by state.

## Introduction

In the United States, there is substantial regional variation in healthcare expenditure and access.^1^ Here, we conduct an ecological study to determine if state-level healthcare access is associated with trajectories of daily reported COVID-19 cases. We hypothesized that differences in ease of healthcare access could translate into differences in accessing COVID-19 testing, and hence impact the number of reported cases. We measured healthcare access using multiple variables at the state level, such as per capita healthcare spending, percentage of uninsured persons, health professional shortages, number of physicians and nurses per 100,000 population, and adoption of Medicaid expansion, among others. Our focus is on the trajectories of daily reported COVID-19 cases, instead of cumulative cases, as trajectories help us identify trends in how the pandemic naturally develops over time, and study the shapes of the curve in different states.

## Methods

We analyzed state-level daily data on confirmed and probable COVID-19 cases from The New York Times for all 50 US states, based on reports from state and local health agencies.^2^ The data spanned from January 21, 2020 to June 16, 2020. State-level daily case data was adjusted for the state’s population. State-level socio-demographic, clinical and healthcare access variables were compiled from the US Census Bureau’s 2018 American Community Survey,^3^ the State Health Access Data Assistance Center (SHADAC),^4^ the 2018 Behavioral Risk Factor Surveillance System and the Henry J. Kaiser Family Foundation.^5^

We identified clusters, or homogenous groups, of states with distinct COVID-19 case trajectories. We used a cluster analysis approach appropriate for time-series data, allowing us to account for daily case data over this span of 148 days. A partition around mediods approach was used with a dynamic time warping similarity metric.^6^ This similarity metric captures the nonlinear similarity between two time-series based on their shape, and was chosen because it has been shown in the literature to be optimal for clustering of time series data.^7^ It is not sensitive to time shifts, which is helpful as some states may have reported earlier cases than other states. The cluster analysis maximizes similarities in the shape of daily case trajectories between states within a cluster, and maximizes differences in the shape of daily case trajectories between clusters. In order to identify the optimal number of clusters, 2-, 3-, 4-, and 5-cluster models were compared using established cluster validity indices (see Supplementary Materials).

For each identified cluster, we plotted the mean (standard error) of daily recorded COVID-19 cases per 1 million population across time. We then tested for differences in characteristics between clusters, using the nonparametric Kruskal Wallis test for continuous variables, and Fisher’s exact test for categorical variables. For continuous variables, we presented the median and interquartile range (IQR).

We also used separate logistic regression models to determine whether healthcare access variables were associated with cluster membership adjusting for potential confounding variables. Adjusted odds ratios (OR) and 95% confidence intervals (CI) are presented.

## Results

Two clusters of states were identified (**Figure 1**). The high case trajectory cluster showed an initial steep rise, and then gradual decline in case numbers, while the low case trajectory cluster showed a slow and steady increase in case numbers, with the two clusters showing approximately equal daily case load at the end of the analyzed time period. The high case trajectory cluster was comprised of 19 states totaling 37.4% of the US population.

**Figure 1.**
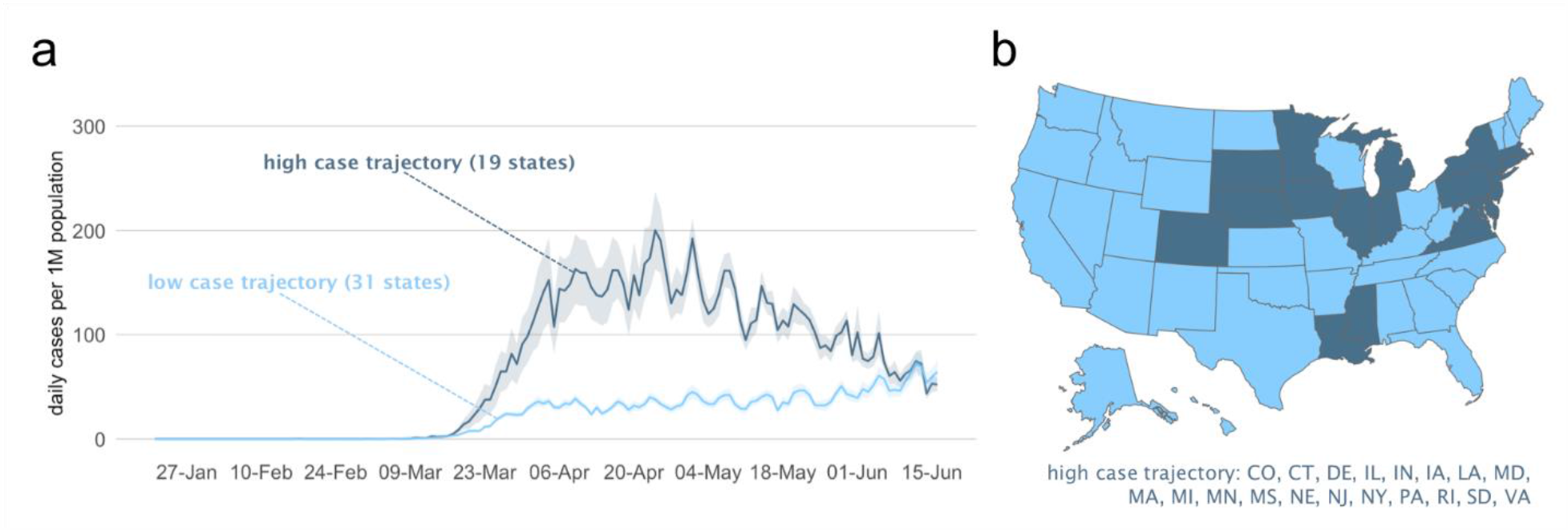
State-level variation in number of daily confirmed and probable cases of COVID-19. **a** Two different trajectories of state-level daily confirmed COVID-19 case numbers, as identified by time-series clustering. Trajectories represent the mean ± standard error of daily confirmed cases, per 1 million (1M) population. **b** Cluster assignment for each state. The higher daily case cluster consisted of 19 states (shaded in dark blue). CO, Colorado; CT, Connecticut; DE, Delaware; IL, Illinois; IN, Indiana; IA, Iowa; LA, Louisiana; MD, Maryland; MA, Massachusetts; MI, Michigan; MN, Minnesota; MS, Mississippi; NE, Nebraska; NJ, New Jersey; NY, New York; PA, Pennsylvania; RI, Rhode Island; SD, South Dakota; VA, Virginia.

**Table 1** compared the two clusters of states by socio-demographic, population health and healthcare access factors. There were significantly more Black residents (p=0.027) and more nursing facility residents (p=0.001) in states reporting high trajectory of COVID-19 cases. States reporting a high trajectory of COVID-19 cases also had fewer uninsured persons (p=0.005), fewer persons who reported having to forgo medical care due to cost (p=0.016), more registered physicians (p=0.002) and more nurses (p=0.03), higher health spending per capita (p=0.01), fewer residents in Health Professional Shortage Areas per 100,000 population (p=0.027), and higher adoption of Medicaid Expansion (p=0.05).

**Table 1:** State-level differences in socio-demographic, population health and healthcare access across the two clusters.

|  | High case trajectory cluster (19 states) | Low case trajectory cluster (31 states) | *p-value* |
| --- | --- | --- | --- |
| Proportion White residents | 0.68 [0.60, 0.78] | 0.74 [0.57, 0.83] | 0.503 |
| Proportion Black residents | 0.10 [0.06, 0.16] | 0.05 [0.02, 0.12] | **0.027** |
| Proportion Hispanic residents | 0.10 [0.06, 0.16] | 0.10 [0.04, 0.13] | 0.623 |
| Proportion Asian residents | 0.04 [0.02, 0.06] | 0.02 [0.02, 0.04] | 0.075 |
| Proportion aged 65 or above | 0.16 [0.16, 0.17] | 0.17 [0.16, 0.18] | 0.216 |
| Proportion in poverty | 0.12 [0.10, 0.13] | 0.13 [0.11, 0.15] | 0.174 |
| Proportion living in urban setting | 0.78 [0.73, 0.88] | 0.70 [0.60, 0.83] | 0.074 |
| Proportion of adults at risk of serious illness if infected with COVID-19 | 0.37 [0.36, 0.40] | 0.39 [0.36, 0.41] | 0.197 |
| Nursing facility residents per 100,000 population | 536 [434, 594] | 388 [251, 469] | **0.001** |
| Proportion with flu vaccination in previous year | 0.37 [0.32, 0.39] | 0.34 [0.31, 0.36] | 0.112 |
| Number of states with no Medicaid expansion | 2 (10.5%) | 12 (38.7%) | **0.05** |
| Proportion uninsured residents | 0.06 [0.05, 0.08] | 0.09 [0.07, 0.11] | **0.005** |
| Proportion residents unable to access medical care due to cost | 0.11 [0.10, 0.12] | 0.13 [0.11, 0.15] | **0.016** |
| Proportion residents who visited a healthcare provider in previous year | 0.76 [0.71, 0.79] | 0.73 [0.69, 0.76] | 0.077 |
| Registered physicians per 100,000 population | 323 [271, 404] | 264 [239, 289] | **0.002** |
| Nurses per 100,000 population | 54 [51, 62] | 49 [41, 57] | **0.03** |
| Health spending per capita (\$) | 8602.00 [8127.50, 9404.50] | 7651.00 [7272.50, 8511.00] | **0.01** |
| Residents in Health Professional Shortage Areas per 100,000 population | 19096 [10964, 26344] | 27646 [19474, 34071] | **0.027** |
| Covid-19 test rate (number tested per 1,000 population) | 77.50 [70.45, 102.05] | 61.70 [48.65, 72.95] | **0.011** |

Next, we analyzed whether healthcare access was associated with cluster membership, after adjusting for proportion of Black residents, proportion of adults at risk of serious illness if infected with COVID-19, proportion of residents residing in urban areas, and number of nursing home facility residents per 100,000. In adjusted models, a higher proportion of uninsured persons (OR: 0.51 [0.25-0.85]; p=0.032), higher proportion of patients who had to forgo medical care due to cost (OR: 0.55 [0.28-0.95]; p=0.048), and no adoption of Medicaid expansion (OR: 0.05 [0 – 0.59]; p=0.04), were associated with reporting a low trajectory of COVID-19 cases.

## Discussion

Our findings show that during the first few months of the COVID-19 pandemic, there was a subset of nineteen states with high daily COVID-19 case trajectories. These states had significantly better healthcare access and more COVID-19 testing. They also had more vulnerable populations, such as higher percentage of Black residents, and more residents living in certified nursing facilities. Our findings from adjusted models suggest that healthcare access can partially explain variations in COVID-19 case trajectories by state.

Our study is limited by potential variation in data quality of reported COVID-19 cases across different states, and there may be other confounding factors that we did not adjust for in our analysis. However, our analysis is strengthened by including a wide range of healthcare access measures, and using the most recent data. Our focus on COVID-19 case trajectories, rather than cumulative cases, allows us to identify similarities and differences in the trend of COVID-19 reported cases over time in different states. Findings from our study may help policymakers better understand the development of the pandemic and, thus, make more informed decisions.

High daily case cluster is comprised of 19 states: Colorado, Connecticut, Delaware, Illinois, Indiana, Iowa, Louisiana, Maryland, Massachusetts, Michigan, Minnesota, Mississippi, Nebraska, New Jersey, New York, Pennsylvania, Rhode Island, South Dakota, Virginia.

P-values calculated using the Kruskal Wallis test for continuous variables and Fisher’s exact test for categorical variables. * denotes significance at the alpha=0.05 level. Unless otherwise specified, values represent median and interquartile range (IQR) for each measure.

No Medicaid Expansion denotes the number of states (%) without adoption of Medicaid Expansion as of February 19, 2020. Physicians and nurses per 100,000 as of March 2019. COVID-19 test rate indicates number of test results per 1,000 population as of June 17, 2020. All other variables are proportions. Poverty defined as below 100% of the federal poverty level. Data sources include the US Census Bureau’s 2018 American Community Survey, the State Health Access Data Assistance Center (SHADAC), the 2018 Behavioral Risk Factor Surveillance System and the Kaiser Family Foundation.

## Data Availability

All data is publicly available from the US Census Bureau 2018 American Community Survey, the State Health Access Data Assistance Center (SHADAC), the 2018 Behavioral Risk Factor Surveillance System and the Henry J. Kaiser Family Foundation. Data on confirmed and probable cases is available from the NYTimes.

https://github.com/nytimes/covid-19-data

## Notes

### Competing Interest Statement

The authors have declared no competing interest.

### Funding Statement

The author(s) received no specific funding for this work.

### Author Declarations

Exempt from IRB. This is an ecological study using data at the state-level. All data is publicly available.

## References

1. 2016 National Healthcare Quality and Disparities Report. Agency for Healthcare Research and Quality; 2016.

2. Coronavirus (Covid-19) data in the United States. The New York Times; 2020. https://github.com/nytimes/covid-19-data. Accessed June 17, 2020.

3. American Community Survey (ACS). United States Census Bureau. https://www.census.gov/programs-surveys/acs.html. Published 2018. Accessed.

4. State health compare. State Health Access Data Assistance Center (SHADAC). http://statehealthcompare.shadac.org/. Accessed June 17, 2020.

5. State data and policy actions to address coronavirus. Kaiser Family Foundation. https://www.kff.org/health-costs/issue-brief/state-data-and-policy-actions-to-address-coronavirus/. Accessed June 17, 2020.

6. Time series clustering along with optimizations for the dynamic time warping distance [computer program]. Version 5.5.6: CRAN R-Project; 2019.

7. Ding H, Trajcevski G, Scheuermann P, Wang X, Keogh E. Querying and mining of time series data: Experimental comparison of representations and distance measures. Proceedings of the VLDB Endowment. 2008;1(2):1542–1552.

